# Subcortical imaging-derived phenotypes are associated with the risk of Parkinson’s disease: A Mendelian Randomization Study

**DOI:** 10.1101/2024.09.24.24314275

**Authors:** Zhichun Chen, Jun Liu, Yong You

**Author notes:** **Correspondence to:** Jun Liu, Department of Neurology and Institute of Neurology, Ruijin Hospital affiliated to Shanghai Jiao Tong University School of Medicine, Shanghai, 200025, China., Yong You, Department of Neurology, The Second Affiliated Hospital of Hainan Medical University, Haikou, 570311, China.

## Abstract

**Background:** The abnormalities of subcortical structures, such as putamen and caudate, play a key role in the occurrence of Parkinson’s disease (PD); however, whether and how imaging-derived phenotypes (IDPs) in subcortical structures are causally associated with the risk of PD remain poorly understood.

**Methods:** The causal associations between subcortical IDPs from UK biobank and risk of PD were evaluated with bidirectional two-sample Mendelian randomization (MR) studies.

**Results:** Totally five subcortical IDPs were found to be causally associated with the risk of PD. Among these IDPs, IDP 168 (Global volume of subcortical gray matter, OR = 1.38 [1.16, 1.63], *P* = 1.82 x 10^−4^), IDP 214 (Right putamen volume, OR = 1.31 [1.15, 1.50], *P* = 7.71 x 10^−5^) and IDP 1441 (T2* signal in right caudate, OR = 1.21 [1.09, 1.35], *P* = 5.23 x 10^−4^) were found to be associated with increased risk of PD. In contrast, IDP 1358 (Mean intensity in right caudate, OR = 0.72 [0.62, 0.85), *P* = 6.77 x 10^−5^) and IDP 1344 (Mean intensity in left caudate, OR = 0.76 [0.65, 0.88], *P* = 3.23 x 10^−4^) were associated with reduced risk of PD.

**Conclusions:** The specific imaging features of the caudate and putamen are causally associated with altered risk of developing PD, thereby providing new insights into the development of novel predictive imaging biomarkers and therapies for PD patients.

## Background

Parkinson’s disease (PD) is the most common movement disorder, affecting millions of individuals all over the world^1^. It is estimated that the global burden of PD has more than doubled due to the increasing numbers of older people, the longer disease duration and environmental factors^2^. It is hypothesized that abnormal deposition of α-synuclein and its pathological propagation between the gut, brainstem, and higher brain structures (i.e., caudate and putamen) probably drive the development and progression of PD^3^. Owing to the insidious onset of disease pathology and clinical symptoms, there is a lack of reliable and sensitive biomarkers for the early prediction of PD^4^. Moreover, currently there is still no cure for PD, so it is urgently needed to develop disease-modifying therapies to delay or even prevent the occurrence of motor and non-motor symptoms in PD patients^5^.

The subcortical structures, especially basal ganglia nuclei, are the key hubs involved in the pathogenesis of PD^3, 6, 7^. For example, altered neural activity in the basal ganglia and thalamus is associated with resting tremor, a core sign of PD^8, 9, 10^. In addition, aberrant neural activity in the basal ganglia is also related to bradykinesia, another cardinal motor sign of PD^11, 12, 13^. Moreover, it is shown that impaired basal ganglia function is associated with the initiation and progression of freezing of gait in PD^14, 15, 16^. Thus, abnormalities of subcortical structures, especially basal ganglia, are associated with the occurrence of key motor signs in PD patients. These findings are supported by a number of magnetic resonance imaging (MRI)-based studies showing imaging-derived phenotypes (IDPs) in subcortical structures are related to clinical manifestations of PD. Particularlly, abnormal functional connectivity in putamen is associated with tremor^17^, cognitive impairment^17, 18^, anxiety disorder^19^, and speech impairment^20^ in PD patients. Anterior caudate atrophy detected by a volumetric study has been shown to be associated with cognitive decline in PD^21^. Moreover, disruption of dorsal stream-caudate structural connectivity is associated with impairment of verbal fluency^22^. Furthermore, transiently reduced activation of caudate nuclei revealed by task-based functional MRI has been shown to be involved in executive dysfunction in early PD^23^. Decreased activation in the left mediodorsal thalamus is associated with depressive symptoms in PD^24^, which is further supported by the finding that impaired white matter microstructure in mediodorsal thalamus is correlated with increased depression severity^25^. PD patients exhibit elevated iron content measured by quantitative susceptibility mapping (QSM) in globus pallidus, putamen, and thalamus compared with control individuals^26^. Importantly, iron accumulation in putamen is associated with worse motor impairment and cognitive decline in patients with PD^27, 28^. Taken together, subcortical IDPs are related to both motor and non-motor symptoms in patients with PD.

As mentioned above, it is generally recognized that IDPs in subcortical structures, such as caudate, putamen, and thalamus, are associated with the clinical features of PD patients; however, whether subcortical IDPs are causally associated with altered risk of PD remains poorly understood. In this study, we determine to investigate the causal relationships between subcortical IDPs and risk of PD using mendelian randomization (MR), which is a powerful methodology utilizing genetic variants as instruments to infer the causal relations between exposure and outcome^29, 30^. Recently, Smith *et al*. (2021) have published the genome-wide associations with 3,935 brain IDPs derived from MRI^31^, thereby providing a valuable source to test the causal relationships between subcortical IDPs and risk of PD with bidirectional MR analysis. Among 3935 brain IDPs, we selected 34 IDPs in several subcortical structures, including caudate, putamen, pallidum, and thalamus, to systematically investigate the causal associations between 34 subcortical IDPs and risk of PD. We hope our study will provide new insights into the early prediction and treatment of PD.

## Methods

### Ethical approval

This MR study was conducted with publicly available summary statistics from previous genome-wide association studies (GWAS), which have been approved by corresponding ethics committee.

### Study design

The study design is shown in Fig. 1. All MR analyses were performed according to standard procedures reported in STROBE-MR checklist^32^ and Burgess *et al*.’s guidelines^33^. In forward MR analysis, each subcortical IDP was treated as the exposure (a total of 34 IDPs; sample size: n = 30056 ∼ 33224)^31^ and the risk of PD (sample size: n = 482,730) was treated as the outcome^34^. In reverse MR analysis, the risk of PD was set as the exposure, with each subcortical IDP as the outcome.

**Fig. 1.**
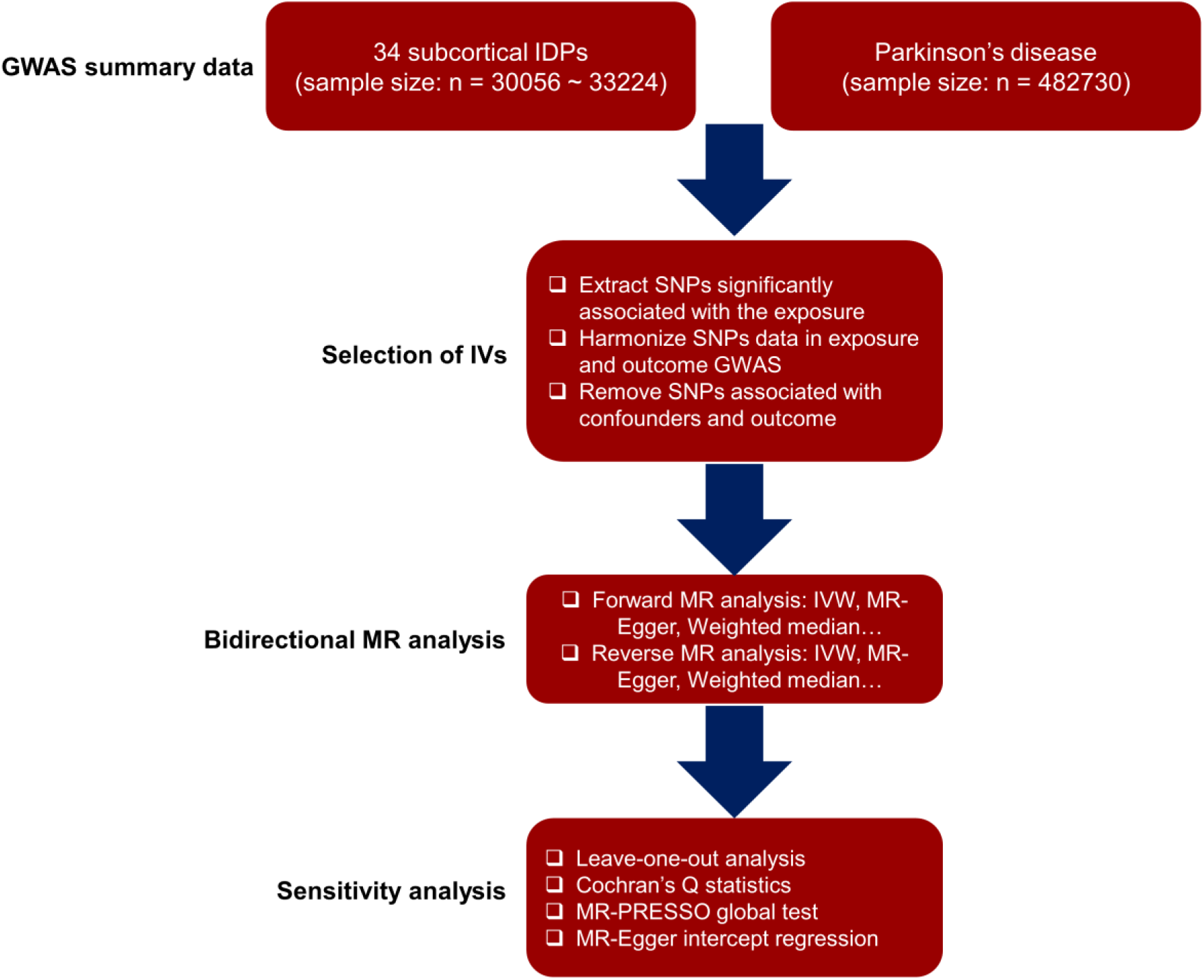
Study design. In the forward MR analysis, each brain IDP was treated as the exposure and the risk of PD as the outcome. In reverse MR analysis, the risk of PD was treated as the exposure and the brain IDP as the outcome. Abbreviations: IDP, Imaging-derived phenotype; GWAS, Genome-wide association study; SNP, single nucleotide polymorphism; IV, Instrumental variable; MR, Mendelian randomization; IVW, Inverse variance weighted; MR-PRESSO, Mendelian Randomization Pleiotropy RESidual Sum and Outlier.

### Exposure data

In forward MR analysis, the GWAS summary statistics of each subcortical IDP were sourced from BIG40 (https://open.win.ox.ac.uk/ukbiobank/big40/)^31^. This dataset consisted of the genome-wide associations of 3935 brain IDPs based on 33,224 European individuals from UK biobank^31^. To identify the causal relationships between subcortical IDPs in both hemispheres and risk of PD, 34 brain IDPs were selected, including 8 subcortical volume and 8 regional intensity metrics (caudate, putamen, pallidum, thalamus) generated by subcortical volumetric segmentation (aseg), 8 subcortical volume metrics derived from T1 images using FMRIB-integrated registration and segmentation tool (FIRST), 8 T2* metrics derived from Susceptibility-weighted imaging (SWI), and 2 global subcortical metrics (global midbrain volume and global subcortical volume). The 34 subcortical IDPs were described in Table S1. In reverse MR analysis, the GWAS summary statistics of PD risk (GWAS ID: ieu-b-7) were extracted from IEU OpenGWAS project (https://gwas.mrcieu.ac.uk/)^34^.

### Outcome data

In forward MR analysis, the GWAS summary statistics of PD risk were sourced from IEU OpenGWAS project (https://gwas.mrcieu.ac.uk/) as mentioned above. In reverse MR analysis, the summary statistics of 34 subcortical IDPs were obtained from IEU OpenGWAS project (https://gwas.mrcieu.ac.uk/).

### Selection of genetic instrumental variables

The instrumental variables (IVs) were selected according to the standard procedures as described previously^29, 30, 32, 33^. Each IV was required to meet 3 assumptions: (i) the IV was significantly associated with the subcortical IDP investigated; (ii) the IVs were mutually independent and not associated with confounding factors; (iii) the IV influenced the risk of PD only through the subcortical IDP. The forward MR analysis used a relaxed statistical *P* threshold (*P* < 1 x 10^−5^) to screen IVs for each subcortical IDP as previously reported^35^. To identify IVs that were independently (r^2^ < 0.001) associated with each subcortical brain IDP, linkage disequilibrium clumping was applied based on 1000 Genomes Project Phase 3 reference panel in a 10 MB window. In the reverse MR analysis, *P* < 5 x 10^−8^ and r^2^ < 0.001 were applied to screen the IVs for the risk of PD. If a particular requested single nucleotide polymorphism (SNP) was not found in the outcome GWAS, then highly correlated proxy SNPs (r^2^ > 0.8) were selected as IVs.

### Removing confounders

PhenoScanner V2 (http://www.phenoscanner.medschl.cam.ac.uk/) and NHGRI-EBI GWAS catalog (https://www.ebi.ac.uk/gwas/docs/file-downloads/) were utilized to remove the SNPs associated with confounders, such as drinking and smoking.

### Two-sample MR analyses

During the MR analysis, multiple statistical methods, including inverse variance weighted (IVW) and weighted median, were performed using TwoSampleMR v0.4.26 R package, while only the IVW method was used as the major statistical approach to examine the causal relationships between each subcortical IDP and the risk of PD due to its high statistical power^36, 37^. Generally, a fixed-effects model was used as the primary causal inference in IVW regression method, whereas a random-effects model was used if the heterogeneity test is significant. In forward MR analysis, MR-RAPS (Robust adjusted profile score)^38^ was specifically performed to assess the statistical robustness of the IVW estimates due to the usage of relatively weak IVs^38^. Bonferroni-corrected *P* < 0.05 (uncorrected *P* < 1.47 x 10^−3^ [0.05/34]) was considered statistically significant. To better interpret the associations between subcortical IDPs and risk of PD, a less stringent false discovery rate (FDR)-corrected *P* < 0.05 was also conducted for error rate control.

### Sensitivity analysis

To ensure the robustness of our findings, a series of sensitivity analyses were conducted. First, Cochran’s Q statistic was employed to examine the heterogeneity of IVs^39^. Secondly, a MR-Egger regression was conducted to detect directional pleiotropy. Thirdly, a MR-PRESSO global test (https://github.com/rondolab/MR-PRESSO/) was performed to test for horizontal pleiotropy. Finally, a leave-one-out (LOO) analysis was conducted to ascertain whether the statistical association was biased by a single IV.

### Data and code availability

GWAS summary statistics of 34 subcortical IDPs were sourced from BIG40 web browser (https://open.win.ox.ac.uk/ukbiobank/big40). All software utilized in the study was publicly accessible, and the download links were provided in the “Methods” section. The code was available to the research community upon reasonable request from the corresponding author.

## Results

### Forward MR: the putative causal effects of subcortical IDPs on the risk of PD

In this bidirectional MR study, we examined the causal associations between subcortical IDPs and the risk of PD. The overall study design was shown in Fig. 1. The initial IVW regression revealed 15 subcortical IDPs may be associated with the risk of PD with FDR method for multiple testing corrections. Among these IDPs, six of them also passed the stringent *P* threshold (*P* < 1.47 x 10^−3^) defined by Bonferroni method (Fig. 2). However, MR estimates of 2 subcortical IDPs (IDP 16 [IDP T1 FIRST right putamen volume] and IDP 1443 [IDP SWI T2star right putamen]) exhibited significant horizontal pleiotropy detected by MR-PRESSO global test (*P* < 0.05). As a consequence, thirteen subcortical IDPs with FDR-corrected *P* < 0.05 passed the sensitivity analysis (Table 1) and 5 subcortical IDPs with Bonferroni-corrected *P* < 0.05 passed the sensitivity analysis (Table 1). As shown in Table 1, Fig. 3 (forest plot), and Fig. 4 (scatter plot), IDP 168 (Global volume of subcortical gray matter, OR = 1.38 [1.16, 1.63], *P* = 1.82 x 10^−4^; Fig. 3A & Fig. 4A), IDP 214 (Right putamen volume, OR = 1.31 [1.15, 1.50], *P* = 7.71 x 10^−5^; Fig. 3B & Fig. 4B) and IDP 1441 (T2* signal in right caudate, OR = 1.21 [1.09, 1.35], *P* = 5.23 x 10^−4^; Fig. 3C & Fig. 4C) were associated with increased risk of PD. By contrast, IDP 1358 (Mean intensity in right caudate, OR = 0.72 [0.62, 0.85), *P* = 6.77 x 10^−5^; Fig. 3D & Fig. 4D) and IDP 1344 (Mean intensity in left caudate, OR = 0.76 [0.65, 0.88], *P* = 3.23 x 10^−4^; Fig. 3E & Fig. 4E) were associated with reduced risk of PD. The LOO analysis supported that the causal effects of 5 subcortical IDPs were not driven by a single SNP (Fig. 5).

**Fig. 2.**
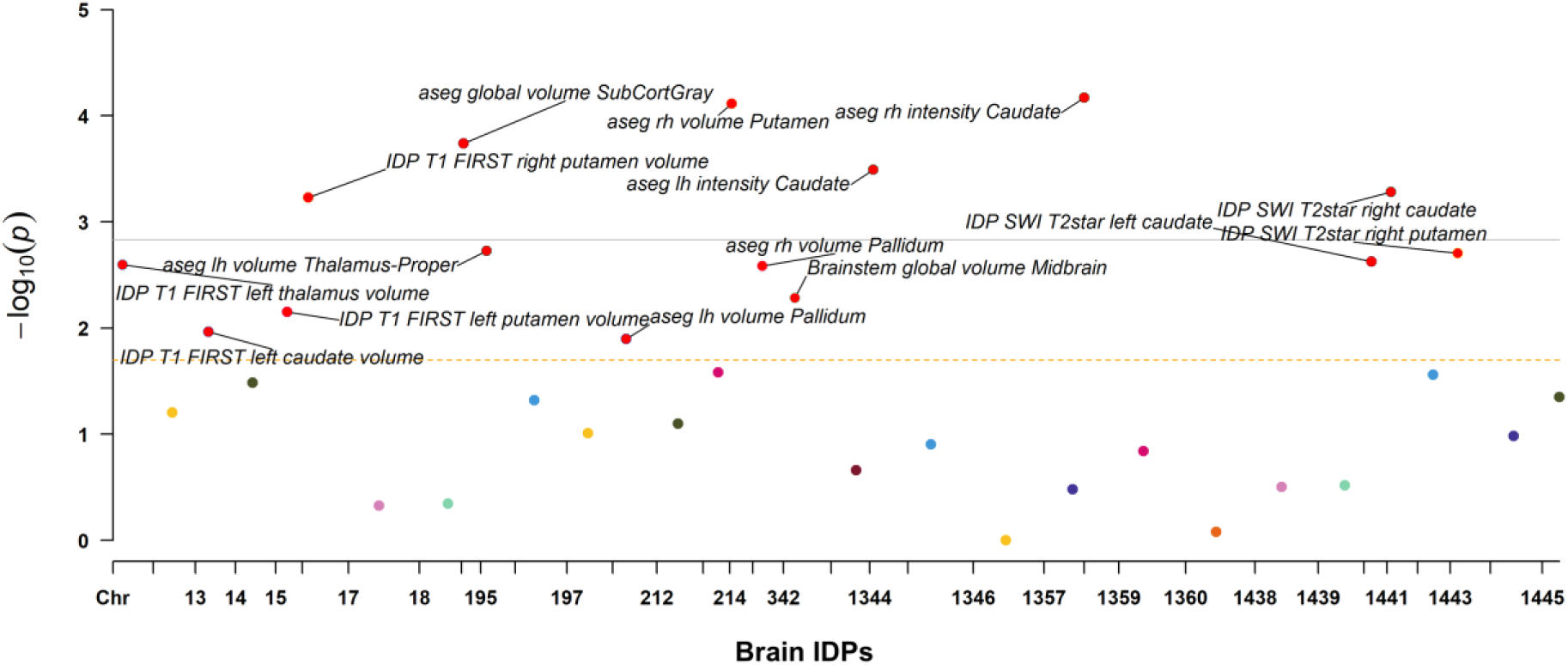
Causal associations between subcortical IDPs and risk of PD in forward MR analysis. Causal effects of subcortical IDPs on the risk of PD were estimated using IVW method. Bonferroni-corrected *P* < 0.05 (uncorrected *P* < 1.47 x 10^−3^ [0.05/34]) was considered statistically significant (Solid line). FDR-corrected *P* < 0.05 was suggested to have potential significance (Dashed line). Abbreviations: IDP, Imaging-derived phenotype; SWI, Susceptibility-weighted imaging.

**Fig. 3.**
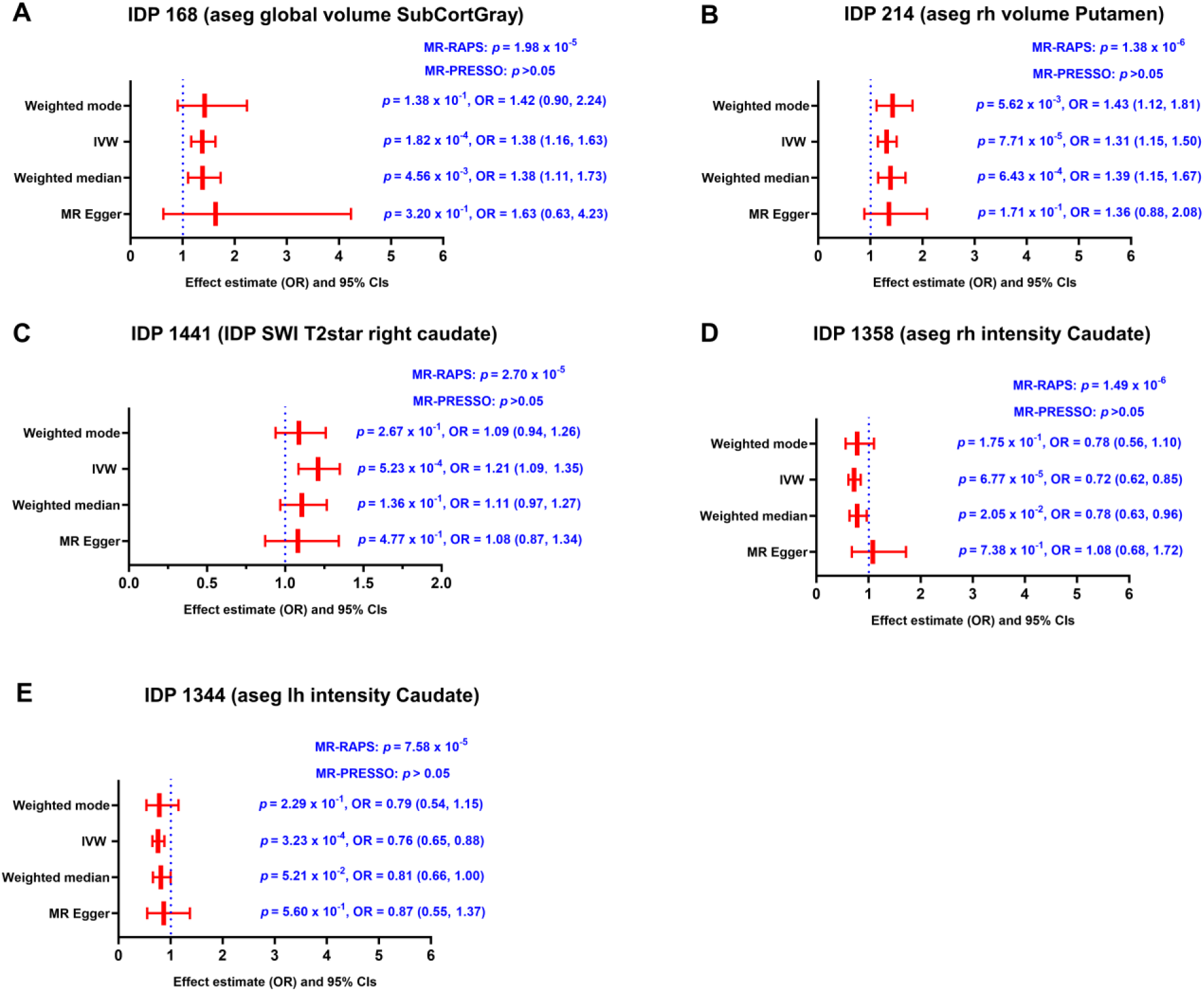
Causalities in the forward MR analysis. **(A-E)** The forest plot shows the significant causalities between IDP 168 (Global volume of subcortical gray matter, **A**), IDP 214 (Right putamen volume, **B**), IDP 1441 (T2* signal in right caudate, **C**), IDP 1358 (Mean intensity in right caudate, **D**), IDP 1344 (Mean intensity in left caudate, **E**) and risk of PD. Causal effects were estimated using four two-sample MR methods (MR-Egger, IVW, weighted median, and weighted mode). MR-RAPS was used to assess the statistical robustness of the MR estimates with relatively weak IVs. MR-PRESSO was used to examine the horizontal pleiotropy. The effect estimates were represented by the OR and 95% CIs. Bonferroni-corrected *P* < 0.05 (uncorrected *P* < 1.47 x 10^−3^ [0.05/34]) was considered statistically significant. Abbreviations: IDP, Imaging-derived phenotype; MR, Mendelian randomization; IVW, Inverse variance weighted; MR-RAPS, Mendelian randomization-robust adjusted profile score; MR-PRESSO, Mendelian Randomization Pleiotropy RESidual Sum and Outlier; OR, Odds ratio; CI: confidence interval.

**Fig. 4.**
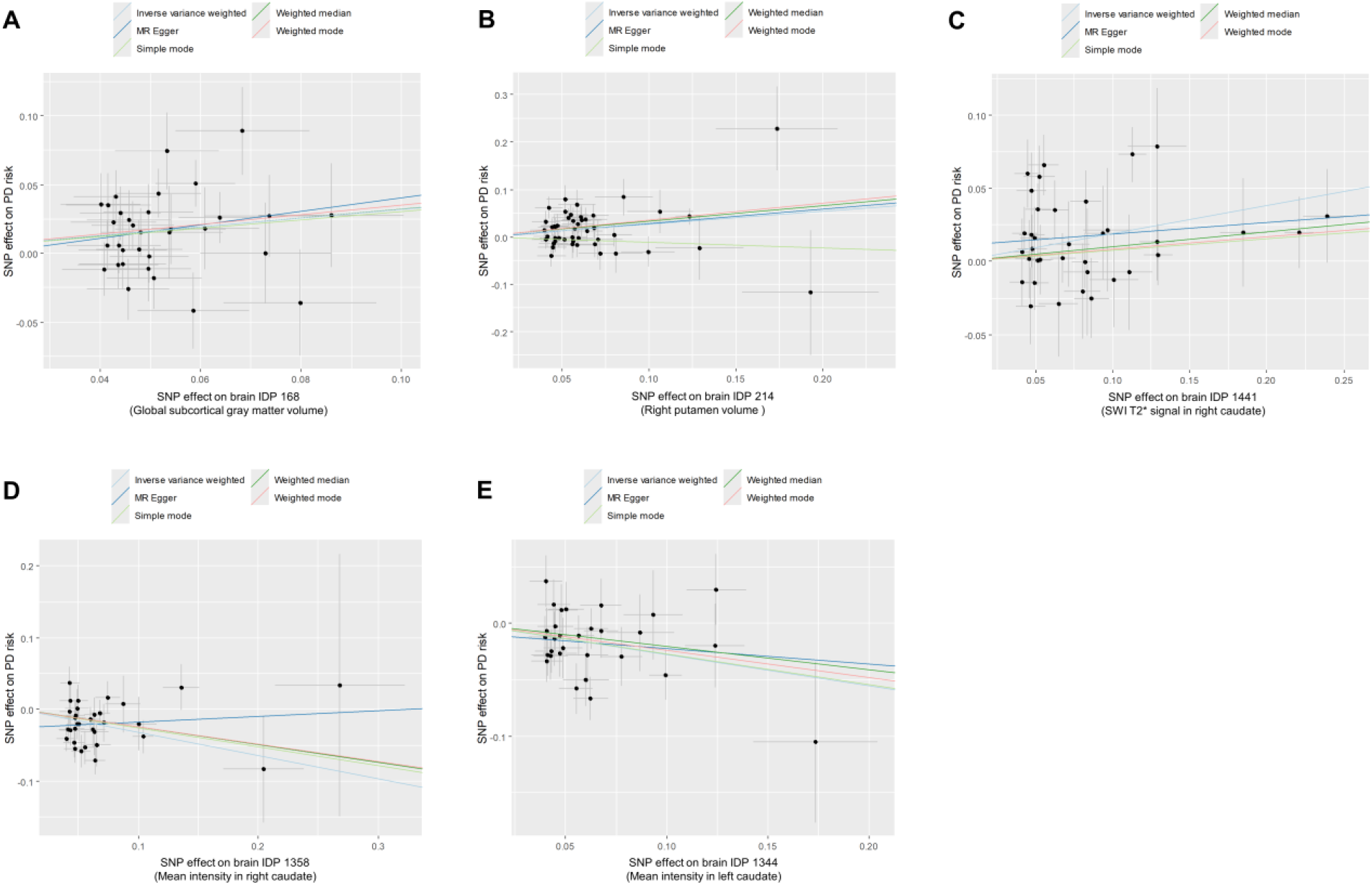
Scatter plots showing the causal effects of subcortical IDPs on the risk of PD. **(A-E)** Scatter plots showing the effects of IDP 168 (Global volume of subcortical gray matter, **A**), IDP 214 (Right putamen volume, **B**), IDP 1441 (T2* signal in right caudate, **C**), IDP 1358 (Mean intensity in right caudate, **D**), and IDP 1344 (Mean intensity in left caudate, **E**) on the risk of PD. Causal effects were estimated using five two-sample MR methods (MR-Egger, IVW, weighted median, weighted mode, and simple mode). Bonferroni-corrected *P* < 0.05 (uncorrected *P* < 1.47 x 10^−4^ [0.05/34]) was considered statistically significant. Abbreviations: PD, Parkinson’s disease; MR, Mendelian randomization; IDP, Imaging-derived phenotype; SNP, Single nucleotide polymorphism.

**Fig. 5.**
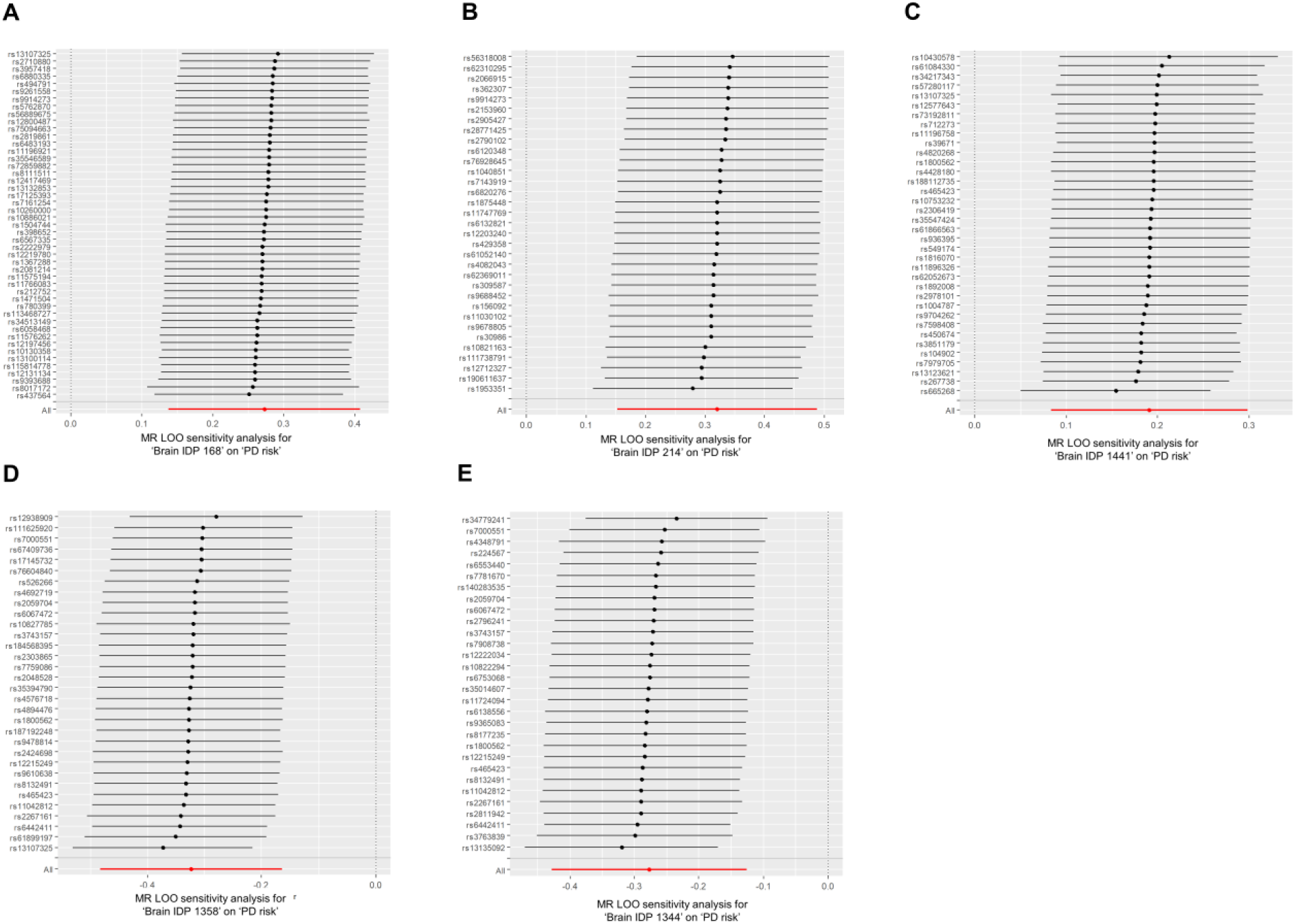
The LOO analysis for significant MR causal estimates. **(A-E)** The LOO analysis for significant MR causal estimates for IDP 168 (Global volume of subcortical gray matter, **A**), IDP 214 (Right putamen volume, **B**), IDP 1441 (T2* signal in right caudate, **C**), IDP 1358 (Mean intensity in right caudate, **D**), and IDP 1344 (Mean intensity in left caudate, **E**). Abbreviations: PD, Parkinson’s disease; IDP, Imaging-derived phenotype; MR, Mendelian randomization; LOO, Leave-one-out.

**Table 1.** The putative effects of subcortical IDPs on the risk of PD examined by forward MR analysis.

| Exposure (IDPs) | Outcome | OR (95%CI) <sup>a</sup> | *P*-value | IV number | Sensitivity analysis <sup>d</sup> | Reverse effect <sup>e</sup> |
| --- | --- | --- | --- | --- | --- | --- |
| aseg rh intensity Caudate<sup>b</sup> | PD | 0.72(0.62, 0.85) | 6.77 x 10<sup>-5</sup> | 32 | passed | No |
| aseg rh volume Putamen<sup>b</sup> | PD | 1.31(1.15, 1.50) | 7.71 x 10<sup>-5</sup> | 47 | passed | No |
| aseg global volume SubCortGray<sup>b</sup> | PD | 1.38(1.16, 1.63) | 1.82 x 10<sup>-4</sup> | 33 | passed | No |
| aseg lh intensity Caudate<sup>b</sup> | PD | 0.76(0.65, 0.88) | 3.23 x 10<sup>-4</sup> | 30 | passed | No |
| IDP SWI T2star right caudate<sup>b</sup> | PD | 1.21(1.09, 1.35) | 5.23 x 10<sup>-4</sup> | 36 | passed | No |
| aseg lh volume Thalamus-Proper<sup>c</sup> | PD | 1.33(1.11, 1.60) | 1.88 x 10<sup>-3</sup> | 25 | passed | No |
| IDP SWI T2star left caudate<sup>c</sup> | PD | 1.17(1.06, 1.29) | 2.37 x 10<sup>-3</sup> | 31 | passed | No |
| IDP T1 FIRST left thalamus volume<sup>c</sup> | PD | 1.34(1.11, 1.61) | 2.53 x 10<sup>-3</sup> | 24 | passed | No |
| aseg rh volume Pallidum<sup>c</sup> | PD | 1.25(1.08, 1.44) | 2.60 x 10<sup>-3</sup> | 36 | passed | No |
| Brainstem global volume Midbrain<sup>c</sup> | PD | 1.21(1.06, 1.38) | 5.18 x 10<sup>-3</sup> | 53 | passed | No |
| IDP T1 FIRST left putamen volume<sup>c</sup> | PD | 1.28(1.06, 1.52) | 7.02 x 10<sup>-3</sup> | 25 | passed | No |
| IDP T1 FIRST left caudate volume<sup>c</sup> | PD | 1.20(1.04, 1.37) | 1.08 x 10<sup>-2</sup> | 51 | passed | No |
| aseg lh volume Pallidum<sup>c</sup> | PD | 1.28(1.05, 1.55) | 1.26 x 10<sup>-2</sup> | 24 | passed | No |

**Abbreviations:** PD, Parkinson's disease; NDDs: Neurodegenerative diseases; MR: Mendelian randomization; Neurodegenerative diseases; IDP: Imaging-derived phenotypes; OR: Odds ratio.
<sup>a</sup>Inverse variance weighted (IVW) method was selected to be the major method to examine causal associations in MR analysis.
<sup>b</sup>Bonferroni-corrected $P < 0.05$ (uncorrected $P < 1.47 \times 10^{-3}$ [0.05/34]).
<sup>c</sup>FDR-corrected $P < 0.05$ .
<sup>d</sup>The sensitivity analysis included LOO analysis, MR-PRESSO global test, and Pleiotropy test.
<sup>e</sup>The reverse MR analysis was conducted to examine the reverse effect.

### Reverse MR: the putative causal effects of genetically proxied risk of PD on subcortical IDPs

The reverse MR analysis revealed no causal effects of genetically determined risk of PD on 34 subcortical IDPs (FDR-corrected *P* > 0.05).

## Discussion

The critical role of subcortical networks in the pathogenesis of PD has been widely acknowledged within the academic community; however, it is unclear whether there are causal relationships between imaging features of subcortical structures and the risk of PD. In this study, we identified five subcortical IDPs that were found to be causally associated with altered PD risk. Among these IDPs, global subcortical gray matter volume, right putamen volume, and T2* signal in right caudate were found to be associated with higher PD risk, whereas mean tissue intensity in left and right caudate were associated with lower PD risk. These findings contribute to the current understanding of PD pathogenesis, thus providing new insights for the discovery of new imaging biomarkers and imaging-based therapeutic targets.

The genetics of brain IDPs have become a popular research topic in the last decade, and previous GWAS studies has reported the genome-wide associations of more than 4000 brain IDPs^31, 40, 41^. These studies have significantly advanced our understanding of the molecular basis of brain structure and function. Subcortical structures play a key role in the regulation of human behavior, such as movement, consciousness, emotions and learning. Satizabal *et al*. (2019) investigated the common genetic variations associated with the volumes of multiple subcortical structures, including the caudate nucleus, globus pallidus, putamen and thalamus, with GWAS of almost 40,000 individuals from CHARGE, ENIGMA and UK Biobank^42^. They identified 48 significantly genetic loci, which were involved in neurodevelopment, apoptosis, axonal transport, synaptic signaling, and inflammation/infection^42^. Therefore, subcortical brain structures are genetically predisposed^42, 43^. Additionally, according to previous studies, subcortical structures have been shown to be genetically correlated with depression^44^, schizophrenia^45, 46^, and PD^47^. Therefore, subcortical structures might share genetic factors with neuropsychiatric diseases, particularly schizophrenia and PD. Indeed, Garcia-Marin *et al*. (2023) have revealed significant positive genetic correlations between PD and subcortical brain volumes^47^, which were consistent with our findings that subcortical brain volumes were causally associated with the risk of PD. It can be reasonably inferred, therefore, that subcortical brain structures and PD might share a similar molecular basis, which was demonstrated by previous studies showing they were both linked with multiple molecular pathways, including chronic inflammation, the mitophagy, disrupted vesicle-trafficking, calcium-dependent, and autophagic pathways^34, 47^. In our study, we found both global subcortical volume and volumes in multiple subcortical structures (i.e., right putamen) were associated with increased PD risk (Bonferroni-corrected *P* < 0.05), which were consistent with a recent finding that a larger volume of the putamen was related to a higher risk for PD^47^. With a less stringent FDR method for multiple testing corrections, we found larger volumes of other subcortical structures were also associated with higher PD risk, including left thalamus, left caudate, right pallidum, left pallidum, and midbrain (FDR-corrected *P* < 0.05). In light of these findings, we may posit that greater volumes of subcortical structures, particularly the putamen, may be associated with an elevated risk of PD. Here, using IVW regression method, we revealed greater volumes of left and right putamen were both associated with higher PD risk; nevertheless, the IVW estimates of left putamen showed significant horizontal pleiotropy as detected by MR-PRESSO global test (*P* < 0.05). It should be noted here that MR-Egger regression estimates of left putamen revealed no evidence of directional pleiotropy (*P* > 0.05). Therefore, we should be careful in interpreting the causal relationship between left putamen volume and PD risk, taking into account the presence of horizontal pleiotropy.

In PD patients, higher volumes of bilateral putamen have been shown to be associated with increased duration of disease^48^. In addition, greater basal ganglia volume was positively correlated with Unified Parkinson’s Disease Rating Scale part-III scores and bradykinesia^49^. Currently, in this study, we found greater volume of putamen was associated with increased risk of PD. One explanation for these findings is that putamen exhibited accelerated aging with advancing age due to the age-related telomere attrition^50^, which triggers individual-specific gene expression patterns that induce compensatory responses to promote putamen expansion and stress responses to drive the occurrence of PD-related neurodegeneration^47^. Several risk genes actually have been reported to positively affect the volume of putamen and the risk of PD, such as *CRHR1* and *KTN1*^47, 51^. This is supported by a previous study showing that putamen in PD patients exhibited compensatory gene expressions during aging^52^. In addition, a significant upregulation of glial cell line-derived neurotrophic factor (*GDNF*) mRNA levels has been observed in the putamen of PD patients^53^, which might promote the expansion of neural cells^54, 55^. Overall, these explanations remain insufficient and require the presentation of more robust evidence to support them. Further research is necessary to elucidate the molecular mechanisms that underpin the causal associations between the putamen and the risk of PD.

In the reverse MR analysis, we found genetically proxied risk of PD had no causal effects on subcortical IDPs; however, imaging studies in several PD cohorts have reported reduced volumes in several subcortical structures, especially putamen in PD patients^56, 57^. This discrepancy indicates that putamen volume may be affected by aging and other environmental factors, but not only genetic factors^58, 59^. It is noteworthy that the results of our MR study, when considered alongside those of other researchers, suggest that putamen volume may undergo bidirectional alterations in patients with PD. That is, although larger putamen volume increases the risk of PD, putamen volume gradually decreases once the clinical symptoms appear. Further longitudinal studies are required to examine the evolution of putamen volume from the pre-disease stage to the clinically-diagnosed stage.

QSM is an MRI-based technique that enables the quantification of iron in specific tissues. It was reported that older individuals exhibit higher iron concentrations in subcortical regions (i.e., caudate nucleus and putamen) than young individuals^60^. Additionally, PD patients display elevated iron levels as determined by QSM in subcortical structures when compared to age-matched control subjects^26^. It is of particular significance that the accumulation of iron in the putamen is associated with increased severity of motor impairment and deterioration in cognitive function in patients with PD^27, 28^. Recent genetic studies have reported the genome-wide associations of QSM-related brain IDPs in UK biobank^31, 61^, thereby providing us an opportunity to explore the causal associations between T2* signals in subcortical structures and the risk of PD. In this study, we found higher T2* signal in right caudate was associated with increased risk of PD, which is consistent with previous findings showing that higher genetically determined iron levels in the caudate, putamen, and substantia nigra were associated with increased PD risk^62^. Thus, our findings and those of others suggest that higher levels of subcortical iron deposition are associated with increased risk of PD. Actually, iron deposition has been demonstrated to induce the development of neuronal α-synuclein pathology by inducing autophagy dysfunction^63^. Increased nigral iron levels were found to be associated with the selective degeneration of dopaminergic neurons in the substantia nigra^64^. In addition, nigral iron deposition has been shown to influence disease severity by impairing basal ganglia network^65^. When considered collectively, the presence of anomalous iron accumulations within subcortical structures therefore serves as a causal contributor in the development of PD^66^. Further research is needed to elucidate the molecular mechanisms underlying the causal associations between subcortical iron deposition and PD risk. Whether T2* signal in caudate can be a predictive imaging biomarker for PD remains poorly understood, though the iron depositions in substantia nigra and red nucleus have been shown to predict PD-related neurodegeneration^67^. With regard to the therapeutic implications, the iron-chelating agent deferiprone has shown no beneficial efficacy in the context of clinical trials^68, 69^. It was therefore recommended that further exploration of iron-targeting therapies be conducted in future studies.

Tissue intensity derived from subcortical volumetric segmentation of T1 images is relatively less investigated according to previous literature. Here, we found higher tissue intensities in bilateral caudate generated by subcortical volumetric segmentation were causally associated with lower risk of PD. These findings indicated that tissue intensity was an important type of brain IDP with notable clinical implications. Future studies are required to explore whether tissue intensity of bilateral caudate can be a reliable and sensitive imaging biomarker for the prediction of PD.

In conclusion, our findings indicate that characteristic imaging phenotypes of the caudate nucleus and putamen are causally related to the risk of developing PD. This provides new directions and perspectives for the development of novel predictive imaging biomarkers and therapies for PD patients.

## Acknowledgments

This work was supported by grants from National Natural Science Foundation of China (Grant No. 81873778, 82071415, and 82060213) and National Research Center for Translational Medicine at Shanghai, Ruijin Hospital, Shanghai Jiao Tong University School of Medicine (Grant No. NRCTM(SH)-2021-03).

## Financial Disclosures

There are no financial conflicts of interest to disclose.

## Supplementary Information

The online version contains supplementary material.

## Notes

### Competing Interest Statement

The authors have declared no competing interest.

