## Supplementary material for "Subcortical imaging-derived phenotypes are associated with the risk of Parkinson’s disease: A Mendelian Randomization Study": Fig. S1

### Supplementary materials

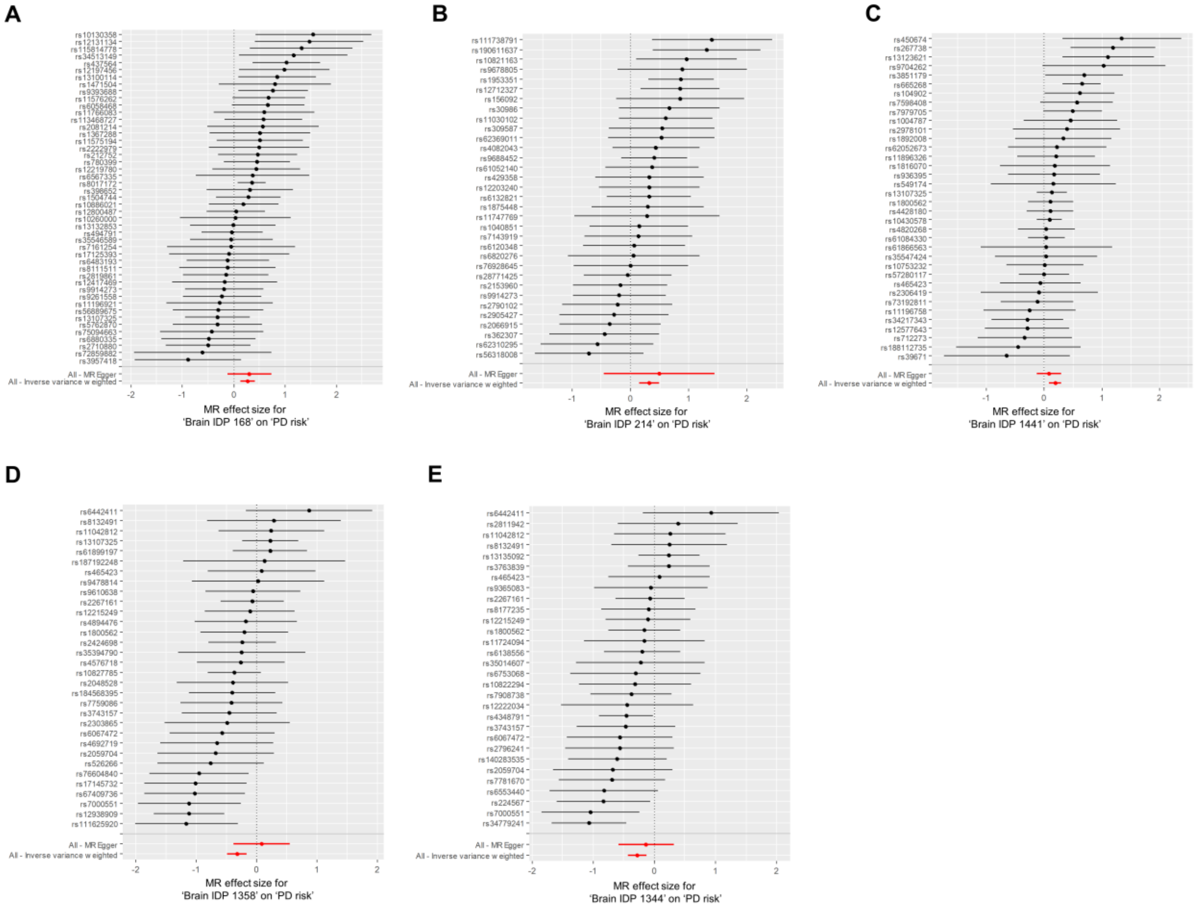


**Fig. S1 The effects of single IV on the risk of PD.** (**A-E**) The forest plots showing the effects of individual IV on the risk of PD. Abbreviations: MR, Mendelian randomization; PD, Parkinson's disease; IDP, Imaging-derived phenotype.
